# Plasma amyloid, phosphorylated tau, and neurofilament light for individualized risk prediction in mild cognitive impairment

**DOI:** 10.1101/2020.07.21.20159129

**Authors:** Nicholas C. Cullen, Antoine Leuzy, Sebastian Palmqvist, Shorena Janelidze, Erik Stomrud, Pedro Pesini, Leticia Sarasa, José Antonio Allué, Nicholas K. Proctor, Henrik Zetterberg, Jeffrey L. Dage, Kaj Blennow, Niklas Mattsson-Carlgren, Oskar Hansson

**Affiliations:** Clinical Memory Research Unit, Department of Clinical Sciences, Lund University, Malmö, Sweden; Araclon Biotech Ltd., Zaragoza, Spain; Eli Lilly and Company, Indianapolis, IN, USA; Department of Psychiatry and Neurochemistry, the Sahlgrenska Academy at the University of Gothenburg, Mölndal, Sweden; Clinical Neurochemistry Laboratory, Sahlgrenska University Hospital, Mölndal, Sweden; Department of Neurodegenerative Disease, UCL Institute of Neurology, Queen Square, London, United Kingdom; UK Dementia Research Institute at UCL, London, United Kingdom; Department of Neurology, Skåne University Hospital, Lund, Sweden; Wallenberg Centre for Molecular Medicine, Lund University, Lund, Sweden; Memory Clinic, Skåne University Hospital, Lund, Sweden

**Author notes:** Contributed equally as first authors (Nicholas C. Cullen, Antoine Leuzy). Contributed equally as senior authors (Niklas Mattsson-Carlgren, Oskar Hansson). **Correspondence and requests for materials should be addressed to**, Oskar Hansson, MD, PhD, Memory Clinic, Skåne University Hospital, SE-205 02, Malmö, Sweden, Or, Niklas Mattsson-Carlgren, MD, PhD, Memory Clinic, Skåne University Hospital, SE-205 02, Malmö, Sweden.

## Abstract

We developed models for individualized risk prediction of cognitive decline in mild cognitive impairment (MCI), using plasma biomarkers of β-amyloid (Aβ), tau, and neurodegeneration. We included MCI patients from the Swedish BioFINDER study (n=148) and the Alzheimer’s Disease Neuroimaging Initiative (ADNI; n=86 for model selection; n=425 for prognostic validation). The primary outcomes were longitudinal cognition and conversion to AD dementia, predicted by plasma Aβ42/Aβ40, P-tau181, and neurofilament light (NfL). A model which included P-tau181 and NfL, but not Aβ42/Aβ40, had the best performance (AUC=0.88 for four-year conversion to AD in BioFINDER, validated in ADNI). The prognostic ability of plasma biomarkers was stronger than a basic model of age, sex, education, and baseline cognition and similar to cerebrospinal fluid biomarkers. Plasma biomarkers, in particular P-tau181 and NfL, may be of high value to identify MCI individuals who will progress to AD dementia in clinical trials and in clinical practice.

## Introduction

About 50 million people live with dementia globally, with prevalence expected to double by 2030.^1^ Fifty to seventy percent of all dementia cases are caused by Alzheimer’s disease (AD).^2^ The dementia stage is preceded by mild cognitive impairment (MCI). Accurate prognosis is important in MCI, since it may either lead to cognitive decline and dementia (due to AD or other diseases) or be benign and stable.^3^ If disease-modifying treatments became available for AD,^4^ accurate prognostics may be important to guide treatment in MCI patients.

Even at the MCI stage, key pathological hallmarks of AD can be detected *in vivo* using cerebrospinal fluid (CSF) biomarkers, *e.g*., the ratio of Aβ42 to Aβ40, and tau phosphorylated at threonine-181 (P-tau181),^5, 6^ or positron emission tomography (PET) of Aβ and tau.^7, 8^ However, the use of these technologies is limited due the perceived invasiveness of lumbar punctures and the high cost and low availability of PET imaging. Blood-based biomarkers could overcome these hurdles.

Blood-based biomarkers of Aβ (A), tau (T), and neurodegeneration (N) in AD^9^ include the Aβ42/Aβ40 ratio,^10^ P-tau181^11, 12, 13^ and neurofilament light (NfL),^14, 15^ respectively. Aβ42/Aβ40 and P-tau181 in plasma correlate with Aβ-PET and tau-PET findings, respectively, and can distinguish AD dementia from controls and non-AD neurodegenerative disorders.^10, 11, 12, 13, 16^ Blood-based NfL is associated with cortical atrophy and cognitive decline in AD.^17^ Most studies on blood-based AD biomarkers report findings at the group level. There is a gap in our understanding of how well these biomarkers predict clinical outcomes at the individual patient level and how they compare to more basic prediction models. Individualized assessment has recently been applied using CSF and related imaging biomarkers in MCI.^18, 19^ A similar study is lacking for blood-based biomarkers. It could be of great value for clinical practice and trials to investigate whether plasma ATN biomarkers performs as well as CSF biomarkers, and better than more basic prediction models. We have previously done a study with a multivariate approach to examine plasma biomarkers and the risk of progression from MCI to AD dementia,^11^ but most other studies focused on evaluating the biomarkers individually.^10, 12, 13^ None of these studies, however, applied the ATN classification system,^9^ or systematically aimed to find the best subset of ATN for individualized predictions.

We measured plasma Aβ42/Aβ40, P-tau181 and NfL in patients with MCI from two large cohorts and tested which subset of plasma biomarkers best predicted individual risk for cognitive decline and progression to AD dementia. We compared the prognostic ability of plasma biomarkers to the same biomarkers measured in CSF, as well as to a more basic prediction model and cross-validated our individual-based risk assessment models both within and across cohorts.

## Methods

The study procedures are outlined in Figure 1.

**Figure 1.**
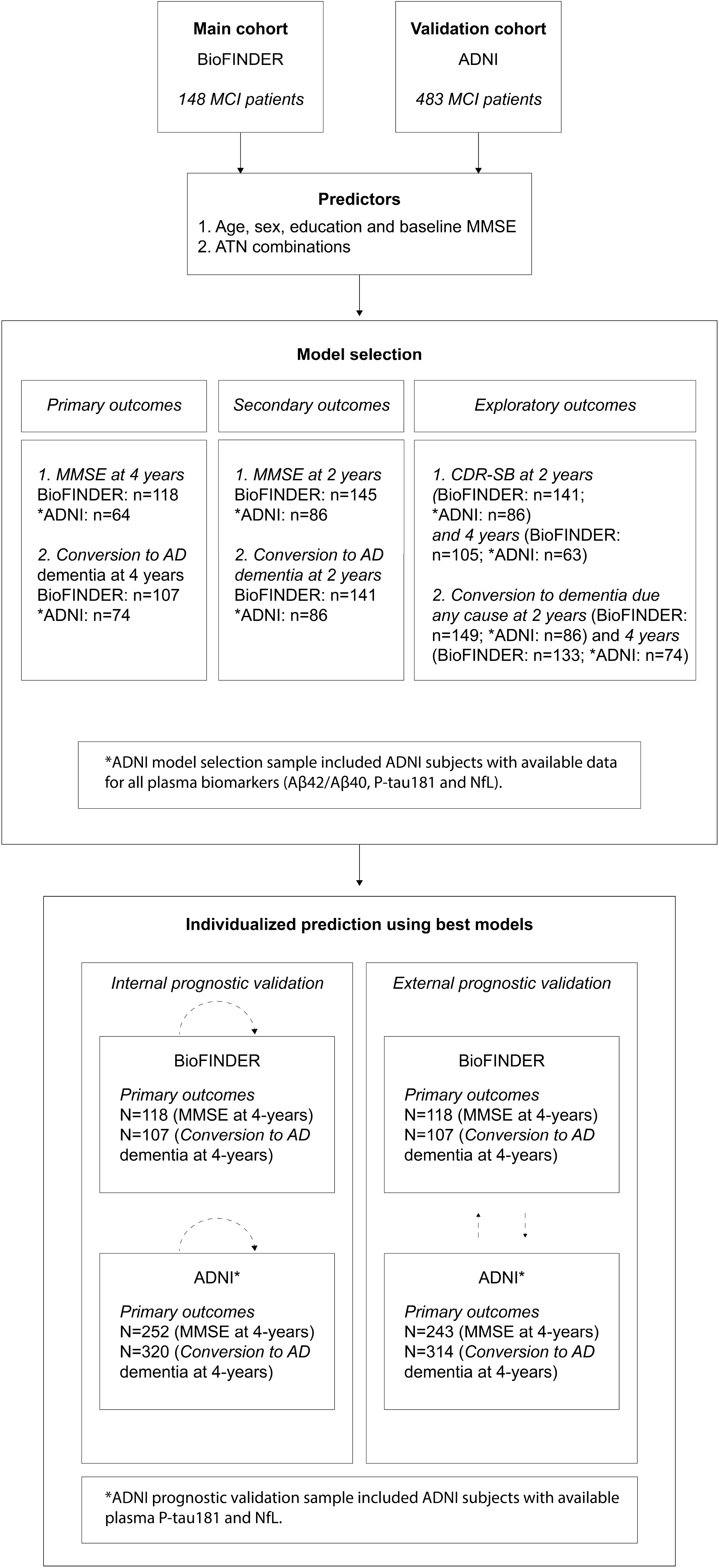
Flow chart of study procedures In ADNI, 425 subjects had data for plasma P-tau181 and NfL; of these 86 also had plasma Aβ42/Aβ40. The number of subjects available for different outcomes differed within both BioFINDER and ADNI. Internal prognostic validation in the section *Individualized prediction using best models* refers to five-fold cross validation for the internal prognostic validation step and validating model selection (i.e. that the best performing model in BioFINDER was the best performing model in ADNI, and vice versa) in the case of external prognostic validation. MCI=mild cognitive impairment; MMSE=Mini Mental State Examination; ATN= Aβ (A), tau (T), and neurodegeneration (N).

### Participants

We studied MCI patients from the Swedish BioFINDER (Biomarkers for Identifying Neurodegenerative Disorders Early and Reliably; clinical trial no. NCT01208675, www.biofinder.se) cohort. The patients were recruited at the memory clinics in the cities of Lund, Malmö and Ängelholm. They were between 60 and 80 years old and fulfilled the consensus criteria for MCI suggested by Petersen et al,^20^ including cognitive complaints, preferably corroborated by an informant; objective cognitive impairment, adjusted for age and education; preservation of general cognitive functioning and a Mini-Mental State Examination score (MMSE) of 24-30; no or minimum impairment of daily life activities, and not fulfilling criteria for dementia, as described previously in detail.^21^ Exclusion criteria included cognitive impairment that could better be accounted for by another non-neurodegenerative condition, severe somatic disease, and current alcohol or substance abuse. After their baseline visit, all patients were seen annually in order to assess clinical progression.

For validation, data were obtained from MCI patients in the Alzheimer’s Disease Neuroimaging Initiative (ADNI) database (adni.loni.usc.edu). The ADNI was launched in 2003 as a public-private partnership, led by Principal Investigator Michael W. Weiner, MD. For up-to-date information, see www.adni-info.org.

All participants gave written informed consent. For BioFINDER, ethical approval was given by the Regional Ethical Committee of Lund University. Ethical approval in ADNI was given by the local ethical committees of all involved sites.

### Outcomes

The co-primary outcomes were the global cognitive measure MMSE and clinical conversion to AD dementia evaluated four years after baseline (“four-year MMSE” and “four-year conversion to AD”, respectively). As secondary outcomes, we used two-year MMSE and two-year conversion to AD dementia. As exploratory outcomes, we used two-year and four-year Clinical Dementia Rating Scale - Sum of Boxes (CDR-SB) along with two-year and four-year conversion to dementia due to any cause.

In BioFINDER, clinical status of dementia due to AD or other diseases was evaluated according to the diagnostic and statistical manual of mental disorders version 5 (DSM-5) criteria for major neurocognitive disorder (*i.e*., dementia). Dementia in ADNI was defined using the National Institute of Neurological and Communicative Disorders and Stroke and the Alzheimer Disease and Related Disorders Association criteria for probable AD.^22^

### Predictors

All models included age, sex, education, and baseline MMSE as predictors (“basic model”). We also measured Aβ42/Aβ40, P-tau181, and NfL in both CSF and plasma.

In BioFINDER, plasma Aβ42/Aβ40 was measured using Elecsys immunoassays on a Cobas e601 analyzer (Roche Diagnostics GmbH, Penzberg, Germany).^10^ A sensitivity analysis was performed using a mass spectrometry-based plasma Aβ42/Aβ40 assay (Araclon Biotech Ltd., Zaragoza, Spain). Plasma P[tau181 was measured on a Meso-Scale Discovery platform (MSD, Rockville, MD), using an assay developed by Eli Lilly.^11^ Plasma NfL was analyzed using a Simoa-based assay.^17^ Moreover, CSF levels of Aβ42 (used in place of Aβ42/Aβ40 due to no available Aβ40 data in ADNI) and P-tau181 were measured using Elecsys assays (Roche Diagnostics GmbH), while CSF NfL was measured using an enzyme-linked immunosorbent assay (UmanDiagnostics AB, Umeå, Sweden).

In ADNI, plasma Aβ42/Aβ40 was analyzed by an immunoprecipitation and mass spectrometry-based method.^23^ P-tau181 was analyzed on a Single molecule array (Simoa) HD-X Analyzer (Quanterix, Billerica, MA), using an assay developed in the Clinical Neurochemistry Laboratory, University of Gothenburg, Sweden.^12^ Plasma NfL was analyzed using the same Simoa-based assay as in BioFINDER.

All biomarker values were natural log transformed. Biomarkers were binarized when validating models across cohorts, whereby cutpoints were defined using Youden’s index to maximize the separation between Aβ-negative cognitively unimpaired (Aβ-CU) participants and Aβ-positive patients with AD dementia (Aβ+ AD) from within each cohort (see Supplementary Text 1); these participants have been described previously.^11, 12^ Note, none of the participants used to define cutpoints were used in the statistical analysis.

### Statistical Analysis

In the first analysis stage (model selection), different linear regression models were fit with cognitive outcomes described above as response variable – a basic model (age, sex, education and baseline MMSE) and plasma biomarker models (the basic model plus different seven different biomarker combinations – Aβ42/Aβ40 only; P-tau181 only; NfL only; Aβ42/Aβ40 and P-tau181; Aβ42/Aβ40 and NfL; P-tau181 and NfL; or all three biomarkers). Models were compared using the coefficient of determination (R^2^) and the Akaike Information Criterion (AIC; lower is better). Statistical significance of different models with the same outcome variable was assessed using the likelihood ratio test. Additionally, logistic regression models were fit with clinical conversion outcomes described above as response variable, with the same set of predictors and the same method of comparison but with area under the curve (AUC) instead of R^2^ as the performance metric.

In the second analysis stage (prognostic validation), the best fitting model identified in the first stage was carried forward and its predictive accuracy was evaluated. Prognostic validation was first done separately within each cohort using 1000 repetitions of five-fold cross validation (internal validation), and then by fitting the model on BioFINDER subjects and testing on ADNI subjects, and vice-versa (external validation; all biomarkers were dichotomized for this analysis, to compare across assays). For internal validation in the BioFINDER cohort, the best fitting plasma model was compared to a CSF model which included CSF Aβ42/Aβ40, P-tau181, and NfL.

In the model selection stage, only participants with all three plasma biomarkers available were used. In the prognostic validation stage, participants were only required to have measurements from the plasma biomarkers included in the best fitting model. Both Q-Q plots and normality of residuals were visually inspected for primary (basic and full ATN) regression models. All analyses were performed using the R programming language (v4.0.0), with significance set at P < .05, two-sided.

## Results

### Study population characteristics – model selection sample

148 MCI patients from the BioFINDER cohort who had all three plasma and CSF biomarker measurements and at least one of the primary or secondary outcomes available were used for model selection (Table 1). The mean age was 71.4 years, mean education was 11.2 years, 36.5% were female, and mean MMSE score was 27.2 ± 1.7 at baseline. Moreover, mean MMSE was 21.8 ± 5.2 four years after baseline and conversion to AD dementia 59.8% within four years of baseline. There was a significant negative correlation between plasma Aβ42/Aβ40 and plasma P-tau181 (R^2^=-0.30, P<0.0001; Supplementary Figure 1) and a significant positive correlation between plasma P-tau181 and plasma NfL (R^2^=0.33, P<0.0001; Supplementary Figure 1), but no significant correlation was observed between plasma Aβ42/Aβ40 and plasma NfL (R^2^=-0.08, P=0.31; Supplementary Figure 1). The associations between corresponding CSF biomarkers in the BioFINDER model selection sample is shown in Supplementary Figure 4.

86 MCI patients from the ADNI cohort who had all three plasma biomarker measurements and at least one of the primary or secondary outcomes available were used to replicate model selection (Table 1). The mean age was 71.5 years, mean education was 16.4 years, 51.2% were female (see Supplementary Text 2 and Supplementary Figure 2 for further description).

**Figure 2.**
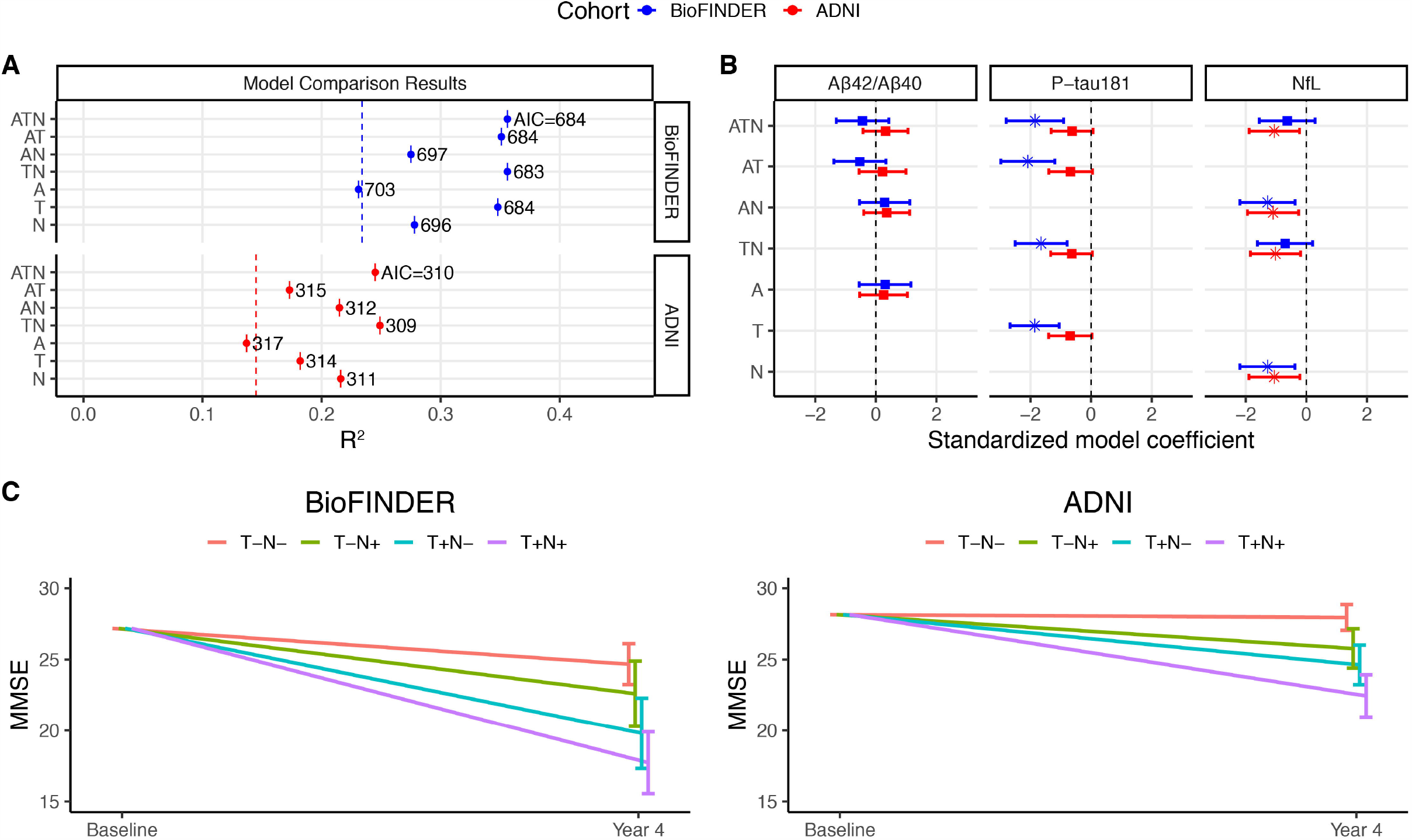
Modelling cognitive decline using plasma Aβ42/Aβ40, P-tau181, and NfL This figure shows the results from modelling cognitive decline in MCI patients using plasma biomarkers. (A) The R^2^ (x-axis) and AIC (values in plot) for each plasma-based model with MMSE evaluated four years after baseline as outcome in ADNI and BioFINDER cohorts are plotted and the basic model performance for reference (dashed line). All models also included age, sex, education and baseline MMSE as predictors. (B) The coefficients from each plasma-based model are shown with MMSE evaluated four years after baseline as outcome in ADNI and BioFINDER cohorts. Statistically significant variables are plotted with an asterisk instead of a square and lines represent 95% confidence intervals. (C) The observed MMSE trajectories (shaded lines) together with the estimated trajectories from the best fitting model (P-tau181 and NfL) according to biomarker status, adjusted for age, sex, education and baseline MMSE.

### Model selection for longitudinal cognition

With four-year MMSE as outcome in the BioFINDER model selection sample (n=118), the model which included plasma Aβ42/Aβ40, P-tau181, and NfL as predictors (“full model”; R^2^=0.36, AIC=684) fit the data significantly better than the basic model (R^2^=0.24, AIC=702, P=0.0001 compared to the full model). However, the best fitting model according to AIC was that which included plasma P-tau181 and NfL, but not Aβ42/Aβ40 (R^2^=0.36, AIC=683). In the best fitting model, there was a significant individual effect of P-tau181 (β=-1.65, P<0.0001) but not NfL (β=-0.70, P=0.13) (Figure 2A-B).

With four-year MMSE as outcome in the ADNI model selection sample used for replication (n=64), the full plasma model (R^2^=0.25, AIC=310) fit the data better than the basic model (R^2^=0.15, AIC=316, P=0.01 compared to full ATN model) and the best fitting model according to AIC again included plasma P-tau181 and NfL only (R^2^=0.25, AIC=309). In the best fitting model, the individual effect of P-tau181 was nearly significant (β=-0.64, P=0.06) while the individual effect of NfL was significant (β=-1.02, P=0.02) (Figure 2A-B).

### Model selection for clinical conversion

With four-year conversion to AD as outcome in the BioFINDER model selection sample (n=107), the full plasma model which included all three biomarkers (AUC=0.88, AIC=106) fit the data significantly better than the basic model (AUC=0.70, AIC=140, P<0.0001 compared to full model). The best fitting model according to AIC included P-tau181 and NfL, but not Aβ42/Aβ40, (AUC=0.88, AIC=104). In the best fitting model, there was a significant individual effect of P-tau181 (OR=5.87, P=0.0001) but not NfL (OR=1.73, P=0.10) (Figure 3A-B).

**Figure 3.**
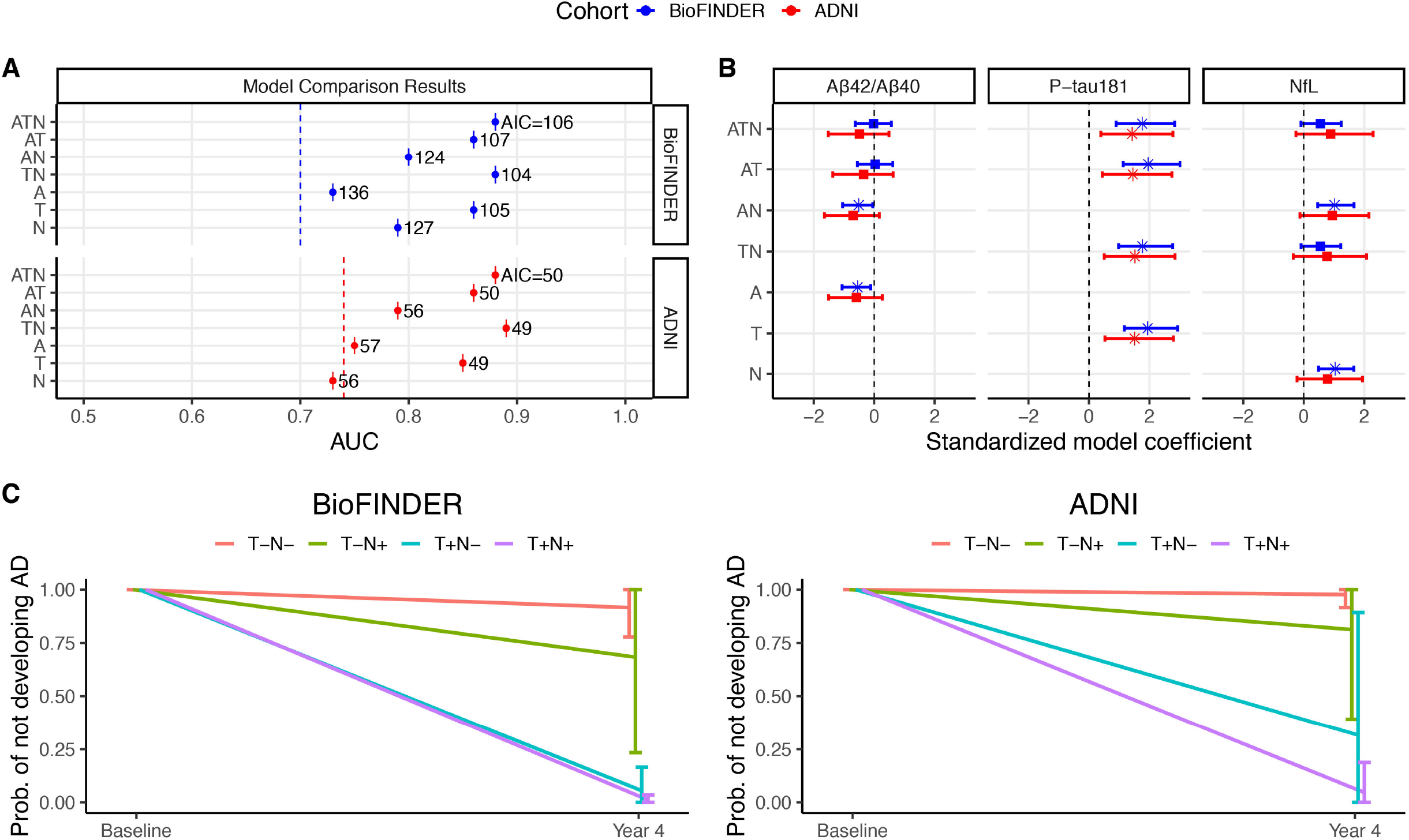
Modelling clinical conversion using plasma Aβ42/Aβ40, P-tau181, and NfL This figure shows the results from modelling clinical conversion in MCI patients using plasma biomarkers. (A) The AUC and AIC values for each plasma-based model with conversion to AD within four years after baseline as outcome in BioFINDER and ADNI cohorts are plotted and the basic model performance is shown for reference (dashed line). All models also included age, sex, education and baseline MMSE as predictors. (B) The coefficients from each plasma-based model are shown with conversion to AD within four years after baseline as outcome in BioFINDER and ADNI cohorts. Statistically significant variables are plotted with an asterisk instead of a square. (C) The estimated probability of not converting to AD as predicted from the best fitting model (P-tau181 and NfL) according to biomarker status, adjusted for age, sex, education and baseline MMSE.

**Figure 4.**
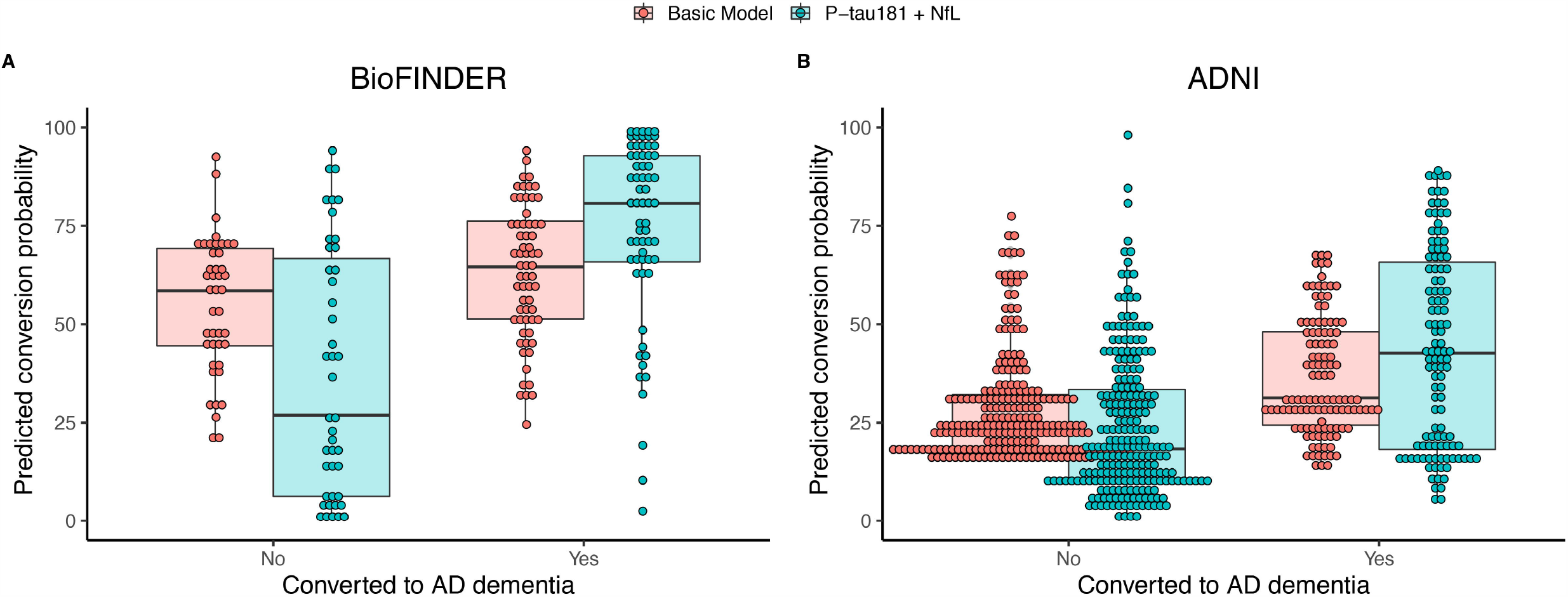
Individualized prediction of conversion from MCI to AD dementia at four-years This figure shows the results from internal cross-validation for clinical conversion for the best performing models as identified in the first stage of analysis using all available BioFINDER (n=107) and ADNI (n=314) subjects. The values plotted here show the predicted probability of conversion from MCI to AD dementia for each individual in the BioFINDER and ADNI cohorts, showing 32.3% improvement (AUC=0.62 for basic model versus AUC=0.82 for P-tau181 and NfL model) of the plasma-based model over of the basic model in BioFINDER and a 15.4% improvement (AUC=0.66 for basic model versus AUC=0.66 for basic model versus AUC=0.76 for P-tau181 and NfL model) in ADNI.

With four-year conversion to AD as outcome in the ADNI model selection sample used for replication (n=74), the full plasma model (AUC=0.88, AIC=50) fit the data better than the basic model (AUC=0.74, AIC=57, P=0.005 compared to full ATN model) and the best fitting model according to AIC again included P-tau181 and NfL only (AUC=0.89, AIC=49). In the best fitting model, the individual effect of P-tau181 was significant (OR=4.58, P=0.009) while the individual effect of NfL was not significant (OR=2.15, P=0.20) (Figure 3A-B).

### Sensitivity analysis using secondary and exploratory outcomes

We performed the same model selection procedure as above using the secondary and exploratory outcomes. We found that the best fitting models identified here varied across outcomes, while the full plasma model including Aβ42/Aβ40, P-tau181 and NfL still always significantly outperformed the basic model alone (Supplementary Tables 1-8).

### Sensitivity analysis using an alternative plasma Aβ42/Aβ40 assay

Because plasma Aβ42/Aβ40 was not selected as part of the best fitting models for the co-primary outcomes, we tested whether this result differed when using a mass spectrometry assay (Araclon Biotech Ltd) instead of the Elecsys assay in the BioFINDER cohort. Here, the best fitting models according to AIC still did not include Aβ42/Aβ40 (Supplementary Tables 7 and 8).

### Study population characteristics – prognostic validation sample

Since the model which included both plasma P-tau181 and NfL (but not Aβ42/Aβ40) provided the best fit across co-primary outcomes in both cohorts, this model was taken forward in the prognostic validation stage. Participants for this analysis were therefore only required to have available plasma P-tau181 and NfL measurements and at least one of the primary or secondary outcomes available. As such, 148 MCI patients from the BioFINDER cohort (no difference from the BioFINDER model selection sample) and 425 MCI patients from the ADNI cohort were included (Table 2). Further description is available in Supplement Table 2.

### Subject-level prognostic validation within cohorts

We performed an internal validation where the subject-level predictive performance of the best fitting plasma model (basic model plus plasma P-tau181 and NfL) was evaluated within each cohort and compared to the basic model alone and to the full CSF model (basic model plus CSF Aβ42/Aβ40, P-tau181, and NfL).

With four-year MMSE as outcome in the BioFINDER prognostic validation sample (n=118), the best fitting plasma model improved cross-validated, out-of-sample prediction compared to the basic model alone (mean absolute error [MAE]=3.07 points versus 3.36 points, P<0.001, 8.5% improvement) and showed no significant difference compared to the full CSF model (P=0.68 over 1000 bootstrapped trials). With four-year MMSE as outcome within the ADNI prognostic validation sample (n=252), the same best fitting plasma model improved out-of-sample prediction compared to the basic model alone (MAE=2.42 points versus MAE=2.49 points, P<0.001, 2.9% improvement) (Figure 4).

With four-year conversion to AD as outcome in the BioFINDER prognostic validation sample (n=107), the best fitting plasma model improved out-of-sample prediction compared to the basic model alone (AUC=0.62 versus AUC=0.82 [note that these cross-validated AUCs are, as expected, lower than the AUCs from corresponding models fit on all data for model selection], P<0.001, 32.3% improvement) and significantly out-performed the full CSF model (P=0.002 over 1000 bootstrapped trials, 5% improvement). With four-year conversion to AD as outcome in the ADNI prognostic validation sample (n=320), the plasma model significantly improved out-of-sample prediction compared to the basic model alone (AUC=0.66 versus 0.76, P<0.001, 15.4% improvement) (Figure 4).

### Subject-level prognostic validation across cohorts

We performed an external validation whereby the subject-level predictive performance of the best fitting plasma model (basic model plus plasma P-tau181 and NfL) was evaluated across each cohort by first fitting the model on BioFINDER subjects and testing on ADNI subjects, and vice-versa. For this analysis, biomarkers were dichotomized.

With four-year MMSE as outcome in the BioFINDER (n=118 prognostic validation sample; of which 28 T-N-, 13 T-N+, 46 T+N-, 31 T+N+) and ADNI (n=243; of which 118 T-N-, 35 T-N+, 46 T+N-, 44 T+N+) prognostic validation samples, the plasma model significantly improved prediction on the test cohort compared to the basic model, both when the model was fit on BioFINDER and tested on ADNI (MAE=3.74 versus 4.08, P=0.0006, 8.3% improvement) and when the model was fit on ADNI and tested on BioFINDER (MAE=4.15 versus 5.19, P<0.0001, 20.1% improvement).

With four-year conversion to AD as outcome in the BioFINDER (n=107; of which 20 T-N-, 5 T-N+, 49 T+N-, 33 T+N+) and ADNI (n=314; of which 139 T-N-, 45 T-N+, 62 T+N-, 68 T+N+) prognostic validation samples, the plasma model improved prediction on the unseen cohort both when the model was fit on ADNI and tested on BioFINDER (AUC=0.76 versus 0.88 [note that these AUCs naturally differ from when models were tested within cohorts, reported in previous sections], P<0.0001, 14.7% improvement) and when the model was fit on BioFINDER and tested on ADNI (AUC=0.77 versus 0.82, P<0.0001, 6.9% improvement).

## Discussion

We addressed the subject-level prognostic value of plasma AD biomarkers (Aβ42/Aβ40, P-tau181 and NfL) in MCI. Plasma P-tau181 in combination with NfL best predicted primary outcomes of decline in MMSE and clinical progression to AD dementia over four years. These results were robust to time horizon (two- or four-years follow-up), selection of outcome (MMSE, CDR-SB, conversion to AD dementia or all-cause dementia), two different cohorts, and choice of Aβ assay. In general, prognostic performance using the plasma-based models were either non-inferior or even better than when using CSF biomarkers, and better than a basic model including age, sex, education and baseline MMSE. These biomarker-driven prediction models can be used in a tool (see panel) for accurate individualized prognosis in MCI; this might improve treatment and care^24^ and could increase power for clinical trials for prodromal AD by only including those with a high risk of future progression.

Our study is novel in the way we address the individualized predictive value of plasma AD biomarkers, but it can be compared to previous work examining CSF and imaging biomarker-driven prognosis at the MCI stage.^19^ Using four separate prognostic models— including age, sex, CSF Aβ42, T-tau and MMSE; and an ATN variant combining CSF Aβ42 and P-tau181 with hippocampal volume—van Maurik and colleagues looked at the likelihood of progression to dementia from MCI.^19^ While all models performed well, the highest performance was seen using the CSF ATN model. Similarly, we found that a combined ATN model (Aβ42/Aβ40, P-tau181 and NfL) in plasma, outperformed a basic model with demographics and baseline MMSE as predictors. Importantly, plasma models improved prediction of longitudinal MMSE despite adjustment for MMSE at baseline, which in itself is very strong predictor of future MMSE.

Inclusion of P-tau181 in the best models may reflect that P-tau181 detects AD-type changes.^11^ On the other hand, plasma Aβ42/Aβ40 was not included in the best model, suggesting that plasma Aβ biomarkers do not provide additional prognostic information in MCI when an efficient plasma P-tau measure is included. This is logical, since symptoms in AD are linked to tau pathology,^25^ and elevations in tau biomarkers appear to be dependent on Aβ pathology.^26, 27^ Findings for plasma Aβ42/Aβ40 have also been more varied than for plasma P-tau181,^10, 28^ and only shown modest reductions in AD dementia (10-15% compared to CU), while P-tau181 is greatly increased in AD dementia (>100% compared to CU).^11, 12, 13^ However, it is possible that plasma Aβ42/Aβ40 may have added value at the preclinical stage of the disease, where it has reached pathological levels,^10^ while tau and neurodegeneration markers continue to increase during the symptomatic stages of the disease.^11, 29^ The best models also included NfL, which is a more general marker of neurodegeneration^17^ and appears to give complimentary prognostic information to P-tau181.

In addition to the CSF studies by van Maurik on individualized biomarker-based risk predictions of dementia in MCI patients,^18, 19^ recent work from our group has examined the association between plasma-based biomarkers and the risk of AD dementia.^11^ Though similar in terms of including plasma Aβ42/Aβ40, P-tau181 and NfL, the present study differs from our previous work in a number of important ways. We now focus on identifying optimal models within the ATN framework, rather than individual biomarkers, and on patient-level (and not group level) predictions. We also compare plasma-based models to a more basic model without biomarkers (but with baseline MMSE) and to CSF-based models. We performed extensive internal and external validation analyses (including novel plasma P-tau181 measurements from ADNI). In terms of results, one important difference compared to the previous study^11^ is the finding that both NfL and P-tau181 (rather than just P-tau181) contribute to the best performing models.

Though the relative importance of biomarkers may vary across contexts and intended applications, plasma biomarkers are promising due to high accessibility and low cost. However, measurement standardization of plasma AD biomarkers will be a considerable task, as it has been for CSF AD biomarkers.^30^

Strengths of our study include the use of CSF-based ATN models as an internal performance benchmark and the focus on risk estimates at the subject level. Validation in two independent cohorts with greatly differing demographic makeup speaks to the robustness and relevance of our findings. BioFINDER patients were recruited in a consecutive fashion at three different memory clinics, with approximately 90% of these referred by primary care physicians.^21^ ADNI patients were recruited from many different clinics and may be more representative of a highly selected clinical trial population. The results were robust across different assays used to measure plasma P-tau181 in BioFINDER and ADNI. We also performed several sensitivity analyses, including for clinical outcomes and for the method used to measure plasma Aβ42/Aβ40. One limitation of the study is the relatively modest sample size. Further studies on larger and more diverse populations, including in primary care, may result in more precise and generalizable models.

## Conclusions

Plasma-based AD biomarkers can provide patient-level prognostic information in MCI, comparable to CSF biomarkers. Plasma P-tau181 in combination with NfL seems to best predict cognitive decline and clinical progression. Plasma biomarkers of core AD features may aid in individualized risk assessment for MCI patients, which represents a critical step towards accessible precision medicine for cognitive diseases. Standardized assays with universal cutoffs, and replication of the findings in large cohorts are needed.

## Data Availability

Plasma and CSF data from ADNI were downloaded online at https://ida.loni.usc.edu. Anonymized data from the BioFINDER study will be shared by request from a qualified academic investigator for the sole purpose of replicating procedures and results presented in the article and as long as data transfer is in agreement with EU legislation on the general data protection regulation and decisions by the Ethical Review Board of Sweden and Region Skane, which should be regulated in a material transfer agreement. The code used for statistical analyses is available at a public repository.

https://brainapps.shinyapps.io/plasmaatnapp/

## Acknowledgments

Work at the authors’ laboratory at Lund University was supported by the Swedish Research Council, the Wallenberg Center for Molecular Medicine, the Knut and Alice Wallenberg foundation, The Medical Faculty at Lund University, Region Skåne, the Marianne and Marcus Wallenberg foundation, the Strategic Research Area MultiPark (Multidisciplinary Research in Parkinson’s disease) at Lund University, the Swedish Alzheimer Foundation, the Swedish Brain Foundation, the Swedish Medical Association, the Konung Gustaf V:s och Drottning Victorias Frimurarestiftelse, the Bundy Academy, the Skåne University Hospital Foundation, and the Swedish federal government under the ALF agreement. Data collection and sharing for the ADNI part of the study was funded by the Alzheimer’s Disease Neuroimaging Initiative (ADNI) (National Institutes of Health Grant U01 AG024904) and DOD ADNI (Department of Defense award number W81XWH-12-2-0012). ADNI is funded by the National Institute on Aging, the National Institute of Biomedical Imaging and Bioengineering, and through generous contributions from the following: AbbVie, Alzheimer’s Association; Alzheimer’s Drug Discovery Foundation; Araclon Biotech; BioClinica, Inc.; Biogen; Bristol-Myers Squibb Company; CereSpir, Inc.; Cogstate; Eisai Inc.; Elan Pharmaceuticals, Inc.; Eli Lilly and Company; EuroImmun; F. Hoffmann-La Roche Ltd and its affiliated company Genentech, Inc.; Fujirebio; GE Healthcare; IXICO Ltd.; Janssen Alzheimer Immunotherapy Research & Development, LLC.; Johnson & Johnson Pharmaceutical Research & Development LLC.; Lumosity; Lundbeck; Merck & Co., Inc.; Meso Scale Diagnostics, LLC.; NeuroRx Research; Neurotrack Technologies; Novartis Pharmaceuticals Corporation; Pfizer Inc.; Piramal Imaging; Servier; Takeda Pharmaceutical Company; and Transition Therapeutics. The Canadian Institutes of Health Research is providing funds to support ADNI clinical sites in Canada. Private sector contributions are facilitated by the Foundation for the National Institutes of Health (www.fnih.org). The grantee organization is the Northern California Institute for Research and Education, and the study is coordinated by the Alzheimer’s Therapeutic Research Institute at the University of Southern California. ADNI data are disseminated by the Laboratory for Neuro Imaging at the University of Southern California.

## Author contributions

AL, NMC and OH conceived the study. AL and NC performed the statistical analysis. ES, SP and OH recruited participants and collected clinical data. HZ, LS, JA, JLD, KB, NP, PP and SJ were responsible for biochemical analyses. NMC developed an online tool implementing the statistical models. AL, NC, LS, JAA, PP, NMC and OH drafted the initial manuscript. All authors contributed to revision and editing of the manuscript.

## Panel: Online individualized risk prediction tool

We provide an online tool at https://brainapps.shinyapps.io/plasmaatnapp/ where individualized predictions can be done for MMSE, conversion to AD dementia, and CDR-SB at 2 year and 4 year after baseline, in patients with MCI at baseline. The tool allows the user to enter age, sex, baseline cognition (MMSE, CDR-SB), and dichotomous biomarker status for CSF or plasma Aβ42/Aβ40, P-tau181 and NfL. It is also possible to test predictions with sparse models including subsets of biomarkers. For example, for a 70-year-old female with MCI and baseline MMSE of 27, the predicted probability of conversion to AD is 33% (90% prediction interval 23-45%) at 2 years and 69% (56-80%) at 4 years, without biomarker information. If all plasma Aβ42/Aβ40, P-tau181 and NfL are known and negative, the probabilities are 6% (2-20%) at 2 years and 16% (5-38%) at 4 years. If all plasma Aβ42/Aβ40, P-tau181 and NfL are positive, the probabilities change to 43% (25-62%) at 2 years and 92% (77-97%) at 4 years.

## Table legends

**Table 1**. Study participant characteristics – model selection

Data are n (%) or mean (SD). M=male; F=female; -/+ indicates negative (normal) or positive (abnormal) biomarker values. Biomarker concentrations are given as pg/mL (natural log transformed). NA= not applicable due the use of different assays or time points in BioFINDER and ADNI; as a result, continuous plasma biomarker data was not compared across groups. P-values refer to an ANOVA analysis across all three groups. All cause dementia at 4-years that were not AD dementia was n=27 for BioFINDER (n=11 vascular dementia, n=8 dementia with Lewy bodies/Parkinson’s disease dementia, n=2 frontotemporal dementia, n=6 non specified dementia) and n=7 for ADNI (n=1 delirium due to West Nile encephalitis, n=1 dementia with Lewy bodies, n=1dementia due to HIV, n=1 normal pressure hydrocephalus, n=1 Down syndrome, n=1 non specified dementia).

**Table 2**. Study participant characteristics in ADNI – prognostic validation

Subset of the ADNI cohort with plasma P-tau181, and NfL measurements and at least one of primary or secondary outcomes available (prognostic validation). M=male; F=female; -/+ indicates negative (normal) or positive (abnormal) biomarker values. Biomarker concentrations are given as pg/mL (natural log transformed).

## Data availability

Plasma and CSF data from ADNI were downloaded online at https://ida.loni.usc.edu. Anonymized data from the BioFINDER study will be shared by request from a qualified academic investigator for the sole purpose of replicating procedures and results presented in the article and as long as data transfer is in agreement with EU legislation on the general data protection regulation and decisions by the Ethical Review Board of Sweden and Region Skåne, which should be regulated in a material transfer agreement. The code used for statistical analyses is available at a public repository.

## References

1. Alzheimer’s Disease International. World Alzheimer Report 2019.

2. Winblad B, et al. Defeating Alzheimer’s disease and other dementias: a priority for European science and society. Lancet Neurol 15, 455–532 (2016).

3. Burnham SC, et al. The dawn of robust individualised risk models for dementia. Lancet Neurol 18, 985–987 (2019).

4. Abbasi J. Promising Results in 18-Month Analysis of Alzheimer Drug Candidate. JAMA 320, 965 (2018).

5. Buchhave P, Minthon L, Zetterberg H, Wallin AK, Blennow K, Hansson O. Cerebrospinal fluid levels of beta-amyloid 1-42, but not of tau, are fully changed already 5 to 10 years before the onset of Alzheimer dementia. Arch Gen Psychiatry 69, 98–106 (2012).

6. Hansson O, et al. CSF biomarkers of Alzheimer’s disease concord with amyloid-beta PET and predict clinical progression: A study of fully automated immunoassays in BioFINDER and ADNI cohorts. Alzheimers Dement 14, 1470–1481 (2018).

7. Rabinovici GD, et al. Association of Amyloid Positron Emission Tomography With Subsequent Change in Clinical Management Among Medicare Beneficiaries With Mild Cognitive Impairment or Dementia. JAMA 321, 1286–1294 (2019).

8. Ossenkoppele R, et al. Discriminative Accuracy of [18F]flortaucipir Positron Emission Tomography for Alzheimer Disease vs Other Neurodegenerative Disorders. JAMA 320, 1151–1162 (2018).

9. Jack CR, Jr., et al. NIA-AA Research Framework: Toward a biological definition of Alzheimer’s disease. Alzheimers Dement 14, 535–562 (2018).

10. Palmqvist S, et al. Performance of Fully Automated Plasma Assays as Screening Tests for Alzheimer Disease-Related beta-Amyloid Status. JAMA Neurol, (2019).

11. Janelidze S, et al. Plasma P-tau181 in Alzheimer’s disease: relationship to other biomarkers, differential diagnosis, neuropathology and longitudinal progression to Alzheimer’s dementia. Nat Med 26, 379–386 (2020).

12. Karikari TK, et al. Blood phosphorylated tau 181 as a biomarker for Alzheimer’s disease: a diagnostic performance and prediction modelling study using data from four prospective cohorts. Lancet Neurol 19, 422–433 (2020).

13. Thijssen EH, et al. Diagnostic value of plasma phosphorylated tau181 in Alzheimer’s disease and frontotemporal lobar degeneration. Nat Med 26, 387–397 (2020).

14. Zetterberg H. Neurofilament Light: A Dynamic Cross-Disease Fluid Biomarker for Neurodegeneration. Neuron 91, 1–3 (2016).

15. Quiroz YT, et al. Plasma neurofilament light chain in the presenilin 1 E280A autosomal dominant Alzheimer’s disease kindred: a cross-sectional and longitudinal cohort study. Lancet Neurol 19, 513–521 (2020).

16. Mielke MM, et al. Plasma phospho-tau181 increases with Alzheimer’s disease clinical severity and is associated with tau- and amyloid-positron emission tomography. Alzheimers Dement 14, 989–997 (2018).

17. Mattsson N, Cullen NC, Andreasson U, Zetterberg H, Blennow K. Association Between Longitudinal Plasma Neurofilament Light and Neurodegeneration in Patients With Alzheimer Disease. JAMA Neurol 76, 791–799 (2019).

18. van Maurik IS, et al. Interpreting Biomarker Results in Individual Patients With Mild Cognitive Impairment in the Alzheimer’s Biomarkers in Daily Practice (ABIDE) Project. JAMA Neurol 74, 1481–1491 (2017).

19. van Maurik IS, et al. Biomarker-based prognosis for people with mild cognitive impairment (ABIDE): a modelling study. Lancet Neurol 18, 1034–1044 (2019).

20. Petersen RC. Mild cognitive impairment as a diagnostic entity. J Intern Med 256, 183–194 (2004).

21. Petrazzuoli F, Vestberg S, Midlov P, Thulesius H, Stomrud E, Palmqvist S. Brief Cognitive Tests Used in Primary Care Cannot Accurately Differentiate Mild Cognitive Impairment from Subjective Cognitive Decline. J Alzheimers Dis 75, 1191–1201 (2020).

22. McKhann G, Drachman D, Folstein M, Katzman R, Price D, Stadlan EM. Clinical diagnosis of Alzheimer’s disease: report of the NINCDS-ADRDA Work Group under the auspices of Department of Health and Human Services Task Force on Alzheimer’s Disease. Neurology 34, 939–944 (1984).

23. Ovod V, et al. Amyloid beta concentrations and stable isotope labeling kinetics of human plasma specific to central nervous system amyloidosis. Alzheimers Dement 13, 841–849 (2017).

24. Gomersall T, Smith SK, Blewett C,Astell A. ’It’s definitely not Alzheimer’s’: Perceived benefits and drawbacks of a mild cognitive impairment diagnosis. Br J Health Psychol 22, 786–804 (2017).

25. Ossenkoppele R, et al. Associations between tau, Abeta, and cortical thickness with cognition in Alzheimer disease. Neurology 92, e601–e612 (2019).

26. Mattsson-Carlgren N, et al. Abeta deposition is associated with increases in soluble and phosphorylated tau that precede a positive Tau PET in Alzheimer’s disease. Sci Adv 6, eaaz2387 (2020).

27. Leuzy A, et al. Diagnostic Performance of RO948 F 18 Tau Positron Emission Tomography in the Differentiation of Alzheimer Disease From Other Neurodegenerative Disorders. JAMA Neurol, (2020).

28. Doecke JD, et al. Total Abeta42/Abeta40 ratio in plasma predicts amyloid-PET status, independent of clinical AD diagnosis. Neurology 94, e1580–e1591 (2020).

29. Mattsson N, Andreasson U, Zetterberg H, Blennow K, Alzheimer’s Disease Neuroimaging I. Association of Plasma Neurofilament Light With Neurodegeneration in Patients With Alzheimer Disease. JAMA Neurol 74, 557–566 (2017).

30. Kuhlmann J, et al. CSF Abeta1-42 - an excellent but complicated Alzheimer’s biomarker - a route to standardisation. Clin Chim Acta 467, 27–33 (2017).

